# Post-operative day 3 Procalcitonin predicts post-operative infectious complications in pancreatic surgery – A systemic review and updated meta-analysis

**DOI:** 10.1101/2020.09.21.20198994

**Authors:** Bhavin Vasavada, Hardik Patel

## Abstract

**Aim of study:** Aim of this meta-analysis is to evaluate post-operative procalcitonin as a marker to predict post-operative infectious complications after pancreatic surgeries.

**Material and Methods:** Systemic literature search was performed using MEDLINE, EMBASE and to identify studies evaluating the diagnostic accuracy of Procalcitonin (PCT) as a predictor for detecting infectious complications on postoperative days (POD) 3 and 5 following pancreatic surgery. A meta-analysis was performed using random effect model and pooled predictive parameters for POD 3 and 5 were derived. Geometric means were calculated for PCT cut offs.

**Results:** 6 studies included day 3 PCT analysis, 2 studies included both day 3 and day 5 analysis. Total data of 471 patients were derived. 161 patients developed infectious complications. Pooled sensitivity, specificity, pooled area under curve, diagnostic odds ratio (DOR), positive and negative like hood ratio of day 3 PCT were 74%,79%,0.8453, 11.03,3.17 and 0.31 respectively. Pooled sensitivity, specificity, diagnostic odds ratio (DOR), positive and negative like hood ratio of day 5 PCT were 83%,70%,12.91,2.91 and 0.25 respectively. Geometric means for PCT cut off for day 3 and 5 were 0.80 and 0.43.

**Conclusion:** Postoperative procalcitonin particularly day 3 procalcitonin levels predict post-operative infectious complications following pancreatic surgeries.

## Introduction

Pancreatic surgeries (Pancreaticoduodenectomy/ distal pancreatectomy) are the main treatments for various benign and malignant disease of pancreas, duodenum, and ampullary region. [1]. Pancreatic surgeries are still associated with very high morbidity and mortality. [2]. Majority of complications following pancreatic surgeries are infectious complications including pancreatic leaks and fistula. [3]. These complications can affect outcomes and also increase cost for pancreatic surgeries. [4].

C reactive protein (CRP) and procalcitonin are suggested as inflammatory markers for diagnosing infective complications following colorectal and abdominal surgeries. [5-10].

CRP is not considered as a specific marker for infection, as it can rise in any inflammatory condition. [11].

Procalcitonin is now emerging as a useful and specific marker for sepsis and guide to antibiotic treatment. [12]. It is suggested as a useful marker in predicting infectious complications for colorectal surgeries. [5]. How every there are very limited literature available studying post-operative procalcitonin as a marker for post-operative infectious complications following pancreatic surgeries.

### Aim of Study

Aim of this meta-analysis is to evaluate post-operative procalcitonin as a marker to predict post-operative infectious complications after pancreatic surgeries.

## Materials and Methods

### Data collection

Medline (PubMed), Embase and Scopus were searched with key words like “procalcitonin” AND “pancreatic surgery”, “pancreaticoduodenectomy”, “distal pancreatectomy”, “post-operative complications”, “infective complication”, “pancreatic leak”, “pancreatic fistula”, “anastomotic leak”. Studies after. Year 2006 were searched. Anastomotic leak and pancreatic fistula were considered as infectious complications and were included in search strategy.

### Definition of post-operative infectious complications

Infectious complications were defined as any complications like intraabdominal abscess, pancreatic leak, pancreatic fistula, wound complications, urinary tract infection, post-operative pneumonia or adult respiratory distress syndrome. Screening was done by two reviewers (BV and HP) independently at the title, abstract, and full text stages. Any disagreements were discussed between the reviewers before a final decision was made.

### Study selection

#### Inclusion criteria

- Randomized control trials
- Observational cohort study
- Studies which included post-operative procalcitonin level between postoperative day 3 to 5.
- Studies where subject underwent pancreaticoduodenectomy or distal pancreatectomy
- Studies which included patients with age 18 and above.
- Studies which evaluated post-operative complications.

#### Exclusion criteria

- Studies where full text articles could not be obtained.
- Studies which included only post-operative day 1 or pre-operative procalcitonin level

#### Data extraction

Information on study characteristics including patient population, study duration, follow-up period, index test, and reference standard were extracted from each study. The primary outcome, i.e., diagnostic performance of PCT to detect infectious complications reported as sensitivity (Se), specificity (Sp), positive predictive value (PPV), negative predictive value (NPV), positive likelihood ratio (LR+), and negative likelihood ratio (LR−) at POD 3 and 5, was collected. As anastomotic leakage or pancreatic fistula were considered a subset of infectious complications and expected to account for most cases of IAI in elective colorectal surgery, it was used as the surrogate outcome of interest during data extraction in studies which did not specifically report infectious complications.

Two-by-two contingency tables were constructed to calculate true-positives (TP), false-positives (FP), false-negatives (FN), and true-negatives (TN) using the reported Se, Sp, PPV or NPV. The calculated TP, FP, TN, and FN were pooled into the meta-analysis.

### Risk of bias assessment

The revised Quality Assessment of Diagnostic Accuracy Studies (QUADAS-2) tool developed by the Cochrane Collaboration was used to assess for the risk of bias and applicability of each study. [13]. The tool consists of four key domains, i.e., patient selection, index test, reference standard, and patient flow through the study and timing of tests. Two reviewers (BV and HP) assessed the study quality independently. In case of disagreement, the judgment was discussed among themselves before a final decision. publication bias was assessed with the Deeks test.

### Statistical analysis

The statistical analysis was performed according to the Preferred Report Items for Systematic Reviews and Meta-analysis (PRISMA) statement. [14]. The pooled prevalence of infectious complications with corresponding 95% confidence interval (95% CI) was calculated using random effect model. The pooled PCT cut-off value was derived using geometric mean of the reported PCT cut-off values. [15]. Using a random effect model, the pooled Se, Sp, LR+, LR−, and diagnostic odds ratios (DOR) with corresponding 95% CI were calculated. Symmetrical summary receiver operating characteristic (SROC) curves were also generated. The area under the curve (AUC) and Q* index (the point on the SROC curve where Se and Sp were equal) were calculated, respectively.[16]. Heterogeneity was assessed using the Higgins I^2^ test, with values of 25, 50, and 75% indicating low, moderate, and high degrees of heterogeneity, respectively. [17]. Meta-regression and subgroup analyses were attempted whenever feasible.

The statistical analysis was performed using Meta-DiSc 1.4 (Hospital Ramon y Cajal and Universidad Complutense de Madrid, Madrid, Spain) and revman 5.4.

## RESULTS

### DATA EXTRACTION

“PUBMED”; “EMBASE”; “SCOPUS” were searched with keywords described as in METHODS. A PRISMA flow chart illustrating the selection of articles for this systematic review is shown in Fig. 1. 127 records identified after above search strategy. (119 from databases, 8 from references of extracted articles). 80 article remained after duplicated removed and screened thoroughly. 69 articles excluded which met exclusion criteria. From the remaining articles 3 excluded as full details were not available. 2 articles excluded as details regarding day 3 and day 5 procalcitonin was not available. Finally 6 articles included in qualitative and quantitative analysis.[18-23].

**Figure 1:**
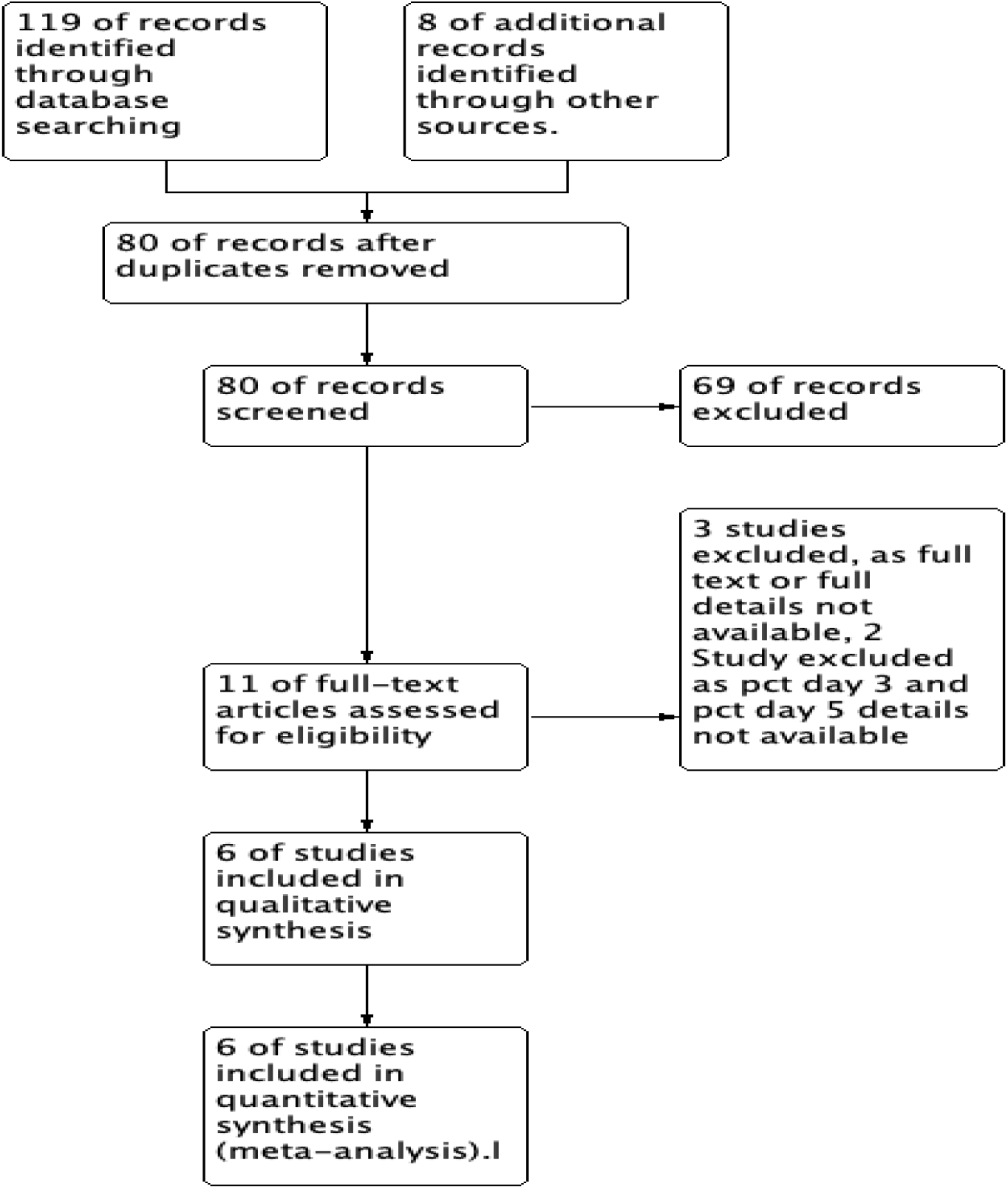
PRISMA FLOW CHART.

### Study characteristics and quality assessment

The characteristics of the 6 studies included in the meta-analysis is described in TABLE 1. All studies included patients who either underwent pancreaticoduodenectomy or distal pancreatectomy. Total 471 patients underwent pancreatic surgery, out of that 161 patients’ developed infectious complications. 452 patients underwent pancreaticoduodenectomy and 19 underwent distal pancreatectomy. All the studies included analysis of Post-operative day 3 procalcitonin analysis. Only 2 studies mentioned post-operative day 5 analysis. We took day 3 and day 5 procalcitonin level as in literature search we could find only one study who analysed day 1 procalcitonin level and also there is risk of high false positive rate due to surgical stress.[24]. Four included studies had infectious complications as their primary outcomes, while two [22,23] had pancreatic leak as its primary outcome.

**TABLE: 1.** STUDY CHARACTERRISTICS.

| METAANALYSIS FINDINGS | POSTOPERATIVE DAY 3 PROCALCITONIN | POSTOPERATIVE DAY 5 PROCALCITONIN |
| --- | --- | --- |
| STUDIES (N) | 6 | 2 |
| NUMBER OF PATIENTS. (N). (INFECTIOUS COMPLICATIONS/TOTAL NUMBER) | 161/471 | 38/105 |
| DERIVED PROCALCITONIN CUTOFF. (NG/ML) [GEOMETRIC MEAN (95% CONFIDENCE INTERVAL] | 0.80 (0.58-1.02) | 0.43 (0.20-0.65) |
| POOLED AUC | 0.8453 |  |
| POOLED SENSITIVITY<br>(%) (95% CI) | 74% (66-81%) | 83% (67-93%) |
| POOLED SPECIFICITY<br>(%) (95% CI) | 79% (74-84) | 70% (58-80) |
| POOLED DOR (95% CI) | 11.03 (4.35-27.96) | 12.91 (3.81-43.7) |
| POOLED POSITIVE LIKE<br>HOOD RATIO (LR+) (95%<br>CI) | 3.17 (2.28-4.40) | 2.91 (2-4.24) |
| POOLED NEGATIVE<br>LIKE HOOD RATIO (LR-)<br>(95% CI) | 0.31 (0.17-0.56) | 0.25 (0.09-0.70) |

### Risk of Bias

The results of the quality assessment using the QUADAS-2 are shown in Figure. 2. Over all there were minimum and unclear concern regarding patient selection in Lida et al. [20] as they excluded surgery where other organs apart from pancreas also needed to be resected and also unclear concern regarding patients flow and timings study by Bianchi et al. [18]. Mintziras et al [23] did not include other complications apart from pancreatic fistula in sensitivity and specificity outcomes.

**FIGURE 2:**
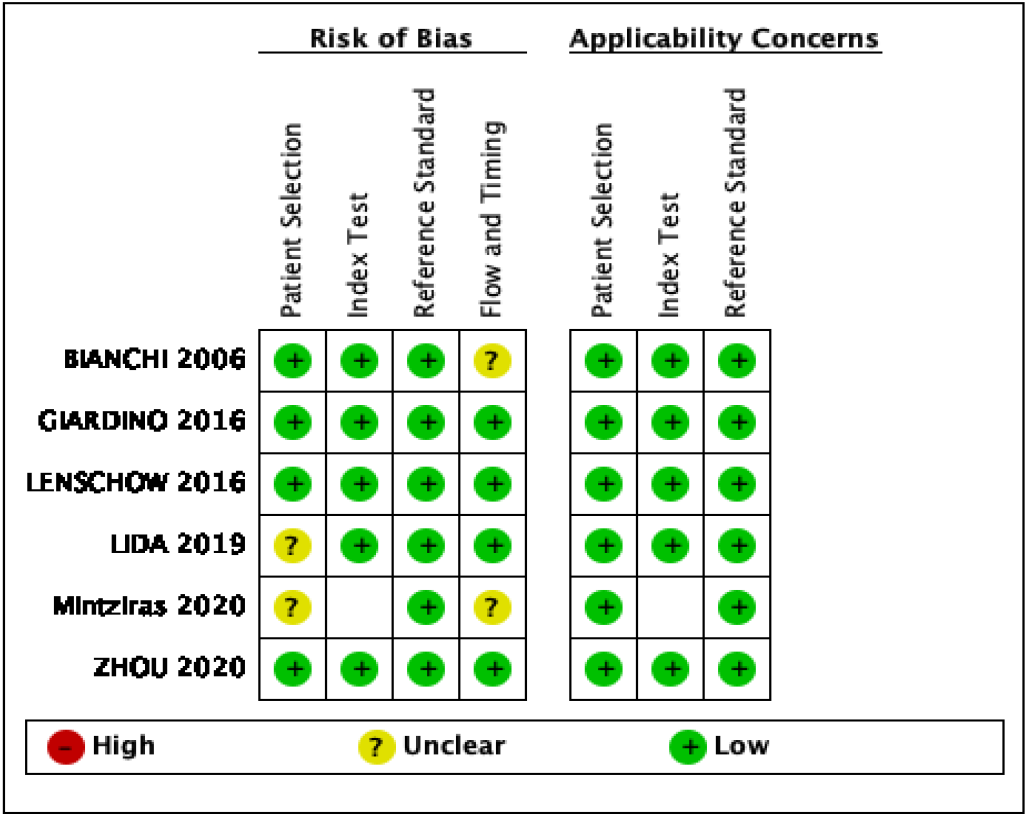
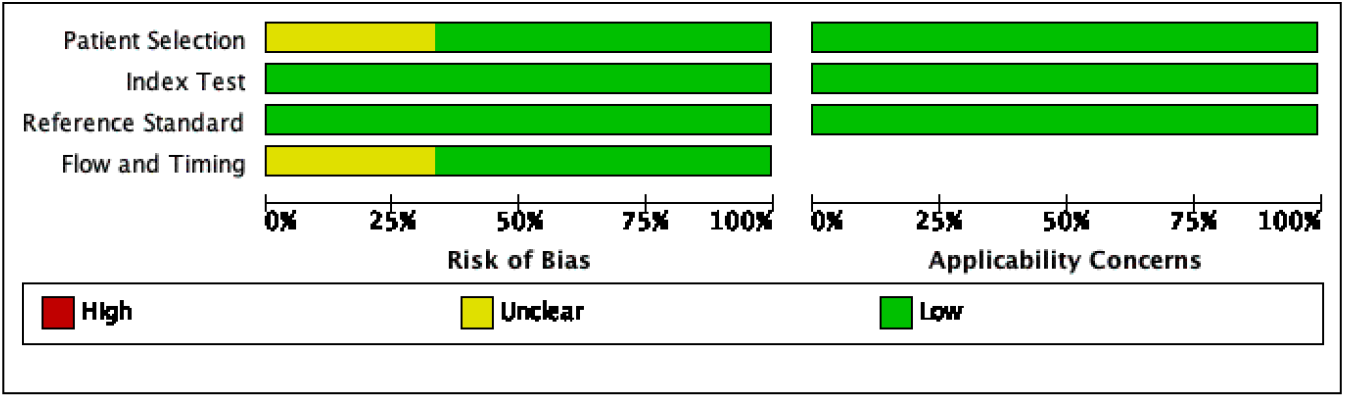
METHODOLOGICAL QUALITY SUMMARY AND GRAPH.

### Predictive value of Procalcitonin for Infectious complications following Pancreatic surgeries

Total 161 patients developed infectious complications out of 471 patients. Overall pooled infectious complication rate after pancreatic surgery is around 34%. A meta-analysis of the predictive value of PCT for infectious complications was performed for POD 3, and 5. The results are summarized in Table 2.

## PROCALCITONIN (PCT) AT POST OPERATIVE DAY 3

All 6 included studies reported day 3 PCT. Geometric mean PCT cut off for predicting infectious complications at day 3 was 0.80 with 95% C.I. 0.58-1.02. Pooled sensitivity of day 3 PCT was 74% with 95% C.I. 66 to 81%. [FIGURE 3]. Pooled specificity was 79% with 95% C.I. 74 to 84%. [figure 4]. SROC curve was prepared which showed pooled Area Under Curve (AUC) 0.8453 with standard error (S.E.) of AUC 0.0394 with Q*= 0.7768 with S.E. (Q)=0.0370. [figure 5]. Positive like hood ratio of day 3 PCT was 3.17 with 95% C.I. 2.28 to 4.40 and negative like hood ratio was 0.31 with 95% C.I. was 0.17-0.56. Pooled diagnostic odds ratio of day 3 PCT in predicting infectious complications was 11.03 with 95% C.I. 4.35-27.96. [Figure 6].Heterogeneity analysis shown with each figure.

**FIGURE 3:**
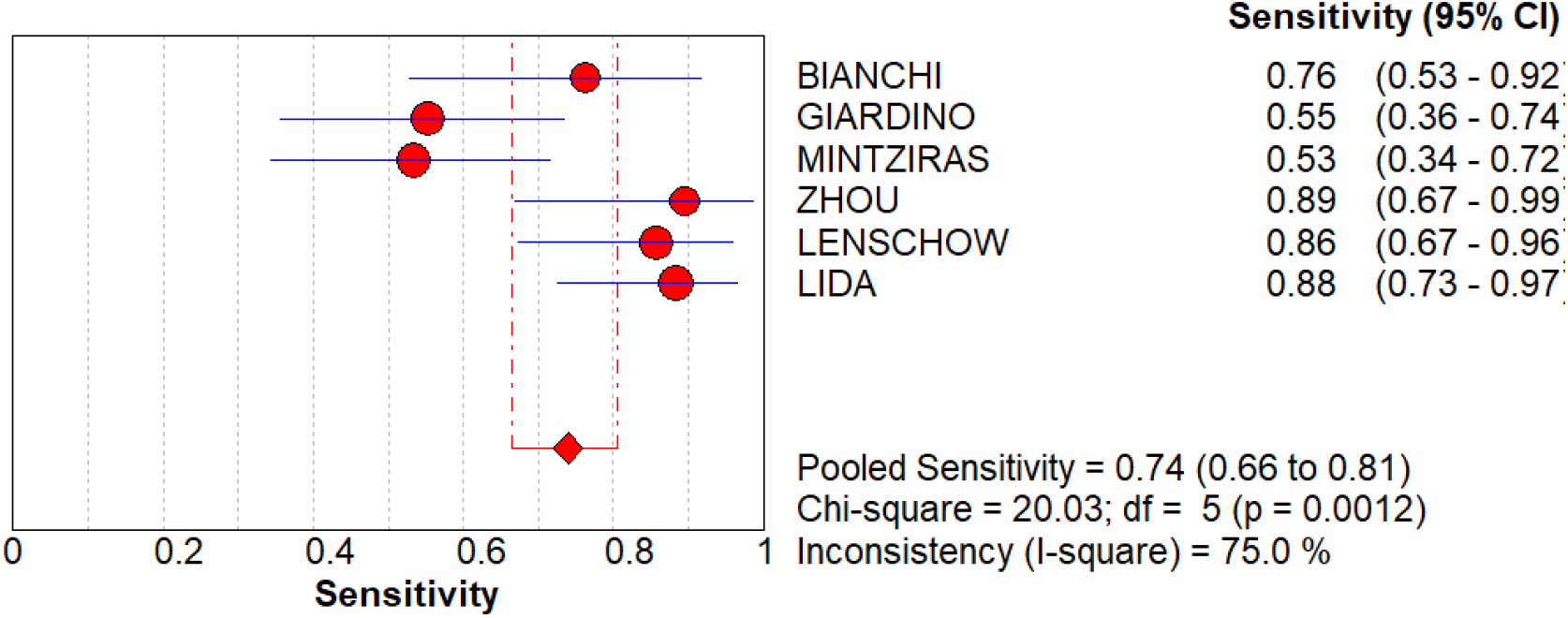
FOREST PLOT OF SENSITIVITY OF DAY 3 PROCALCITONIN IN PREDICTING INFECTIVE COMPLICATIONS.

**FIGURE 4:**
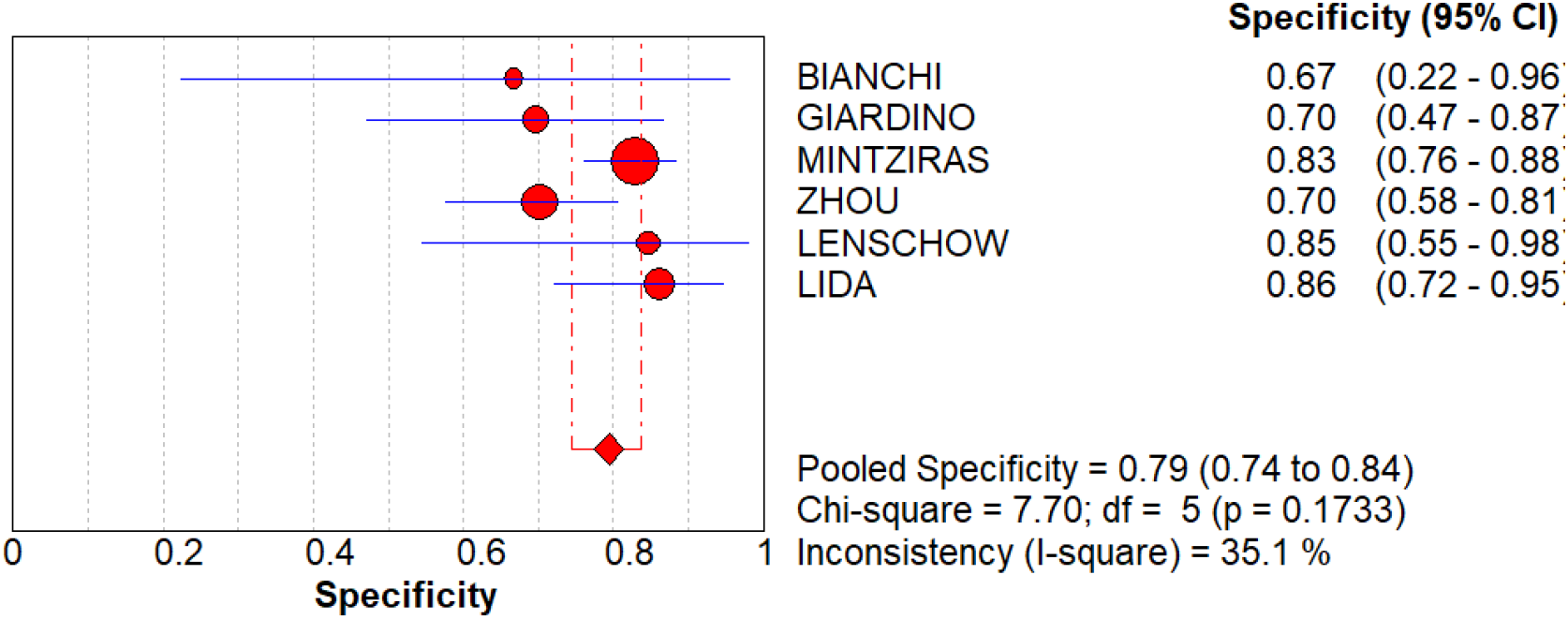
FOREST PLOT OF SPECIFICITY OF DAY 3 PROCALCITONIN IN PREDICTING INFECTIVE COMPLICATIONS.

**FIGURE 5:**
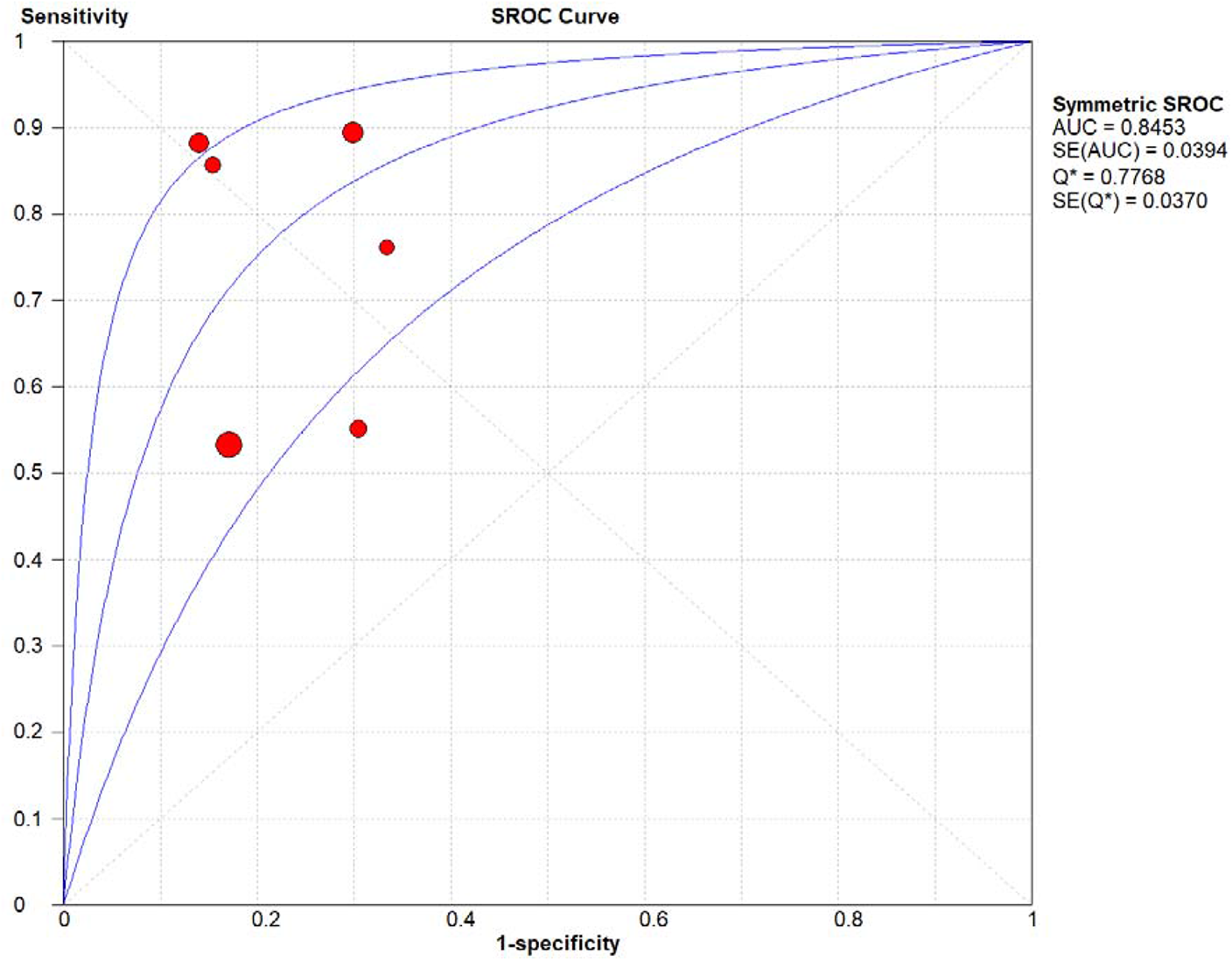
SROC CURVE FOR DAY 3 PROCALCITONIN.

**FIGURE 6:**
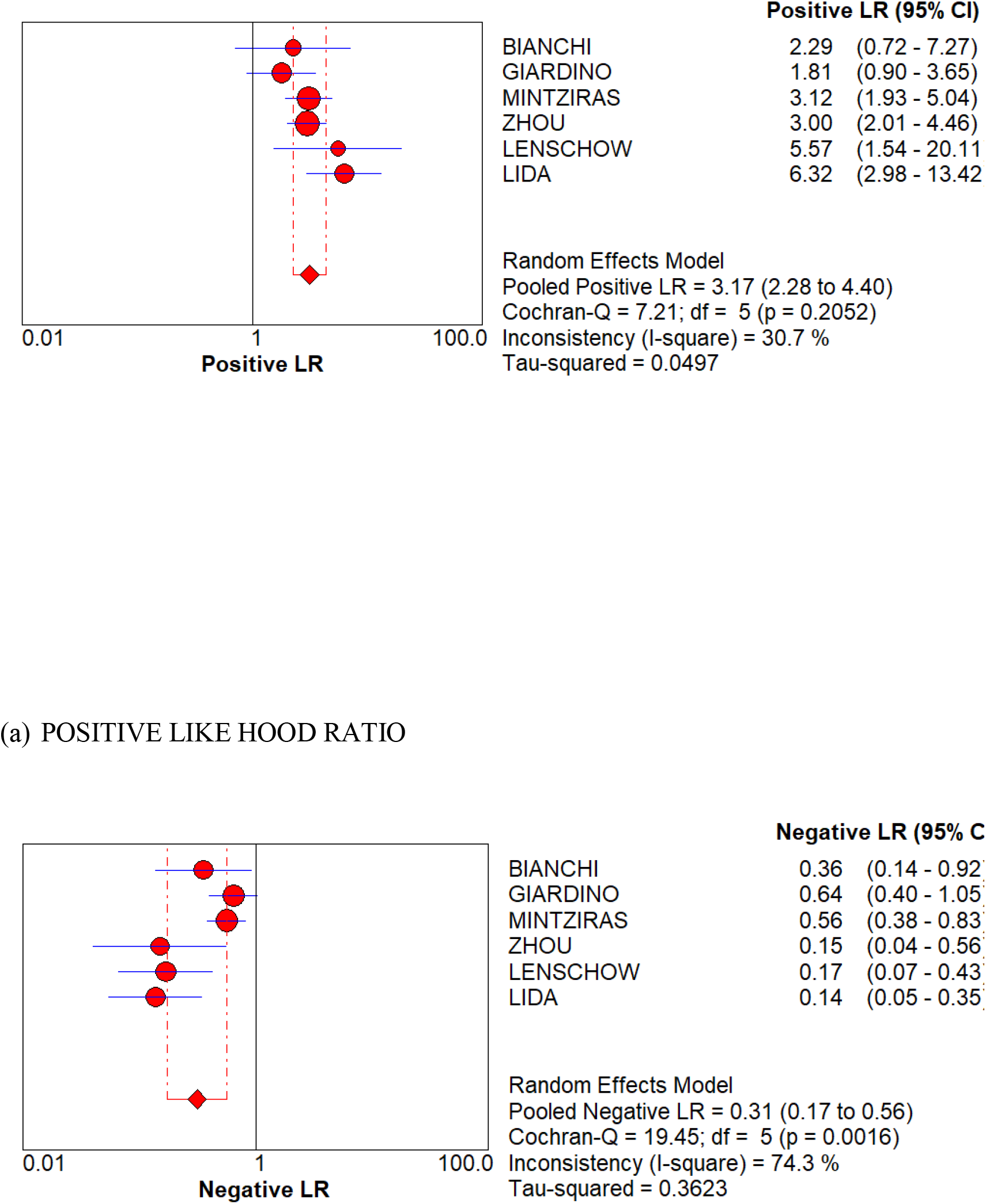
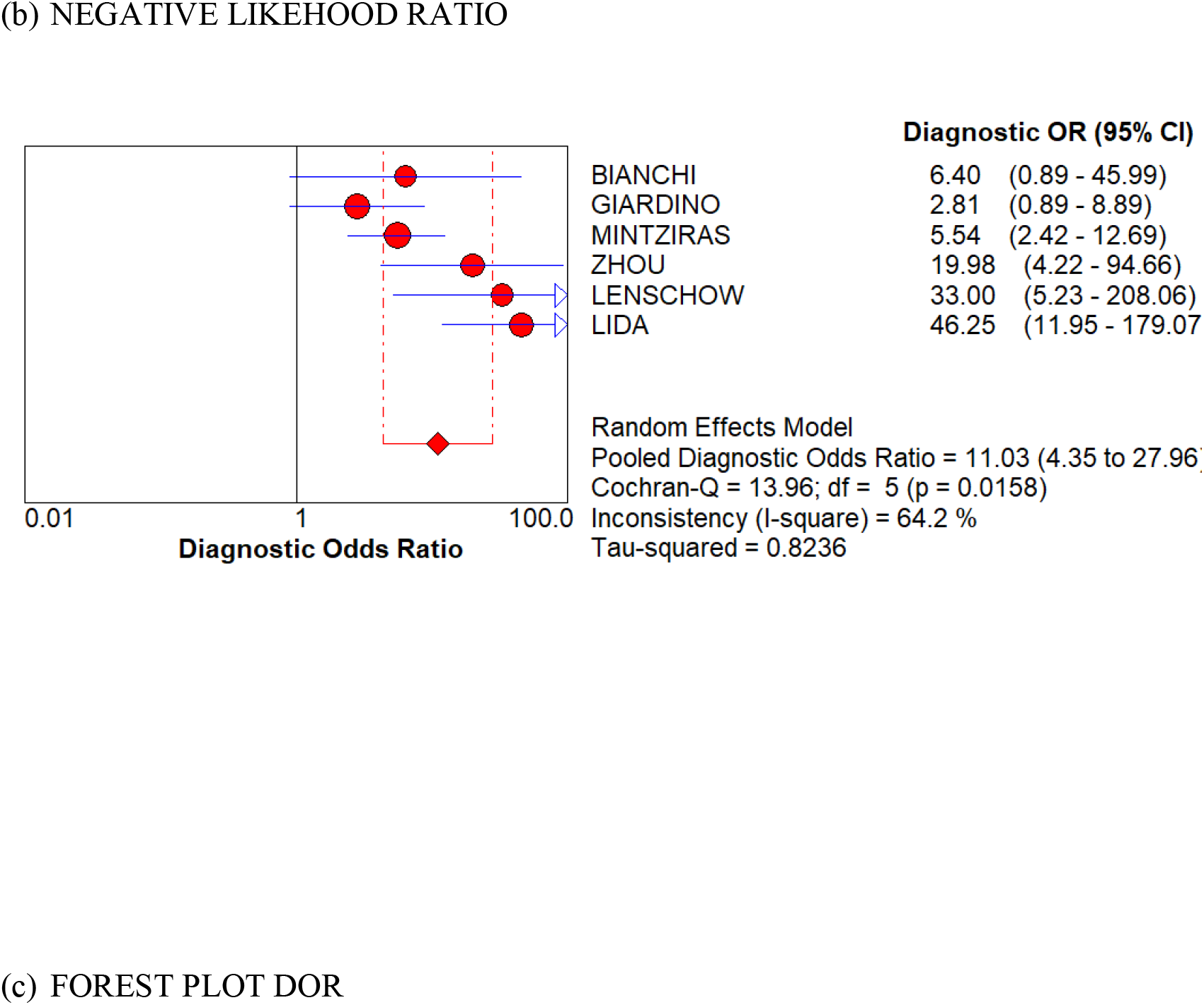
(a) FOREST PLOT FOR POSITIVE LIKE HOOD RATIO (b) FOREST PLOT FOR NEGATIVE LIKEHOOD RATIO. (c) FOREST PLOT DOR.

## PROCALCITONIN (PCT) AT POST OPERATIVE DAY 5

Only 2 studies out of 5 carried out sensitivity and specificity analysis of day 5 PCT. Geometric mean PCT cut off for predicting infectious complications at day 5 was 0.43 with 95% C.I. 0.20-0.65. Pooled sensitivity of day 5 PCT was 83% with 95% C.I. 67 to 93%. Pooled specificity was 70% with 95% C.I. 58 to 80%. [FIGURE 7]. Positive like hood ratio of day 5 PCT was 2.91 with 95% C.I. 2.00 to 4.24 and negative like hood ratio was 0.25 with 95% C.I. was 0.09-0.70. Pooled diagnostic odds ratio of day 5 PCT in predicting infectious complications was 12.91 with 95% C.I. 3.81-43.7. [Figure 8].SROC analysis was not possible as only 2 studies were included however ROC plane with coordinates shown in Figure 9. Heterogeneity analysis shown with each figure.

**FIGURE 7.**
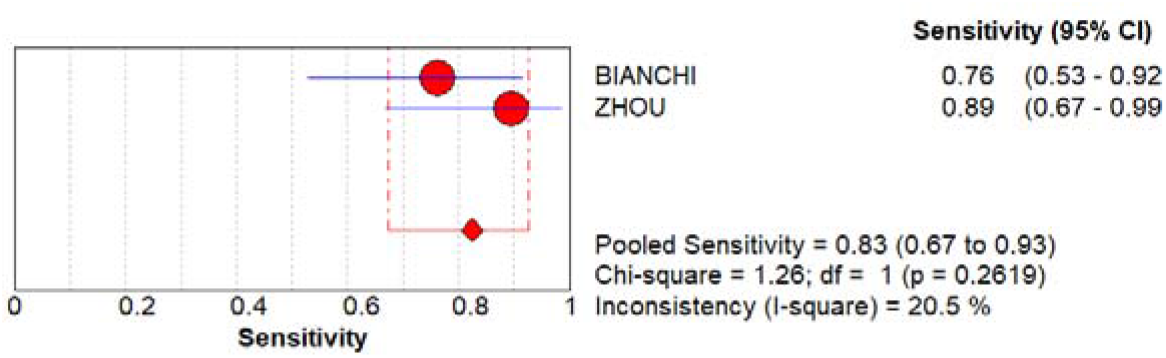
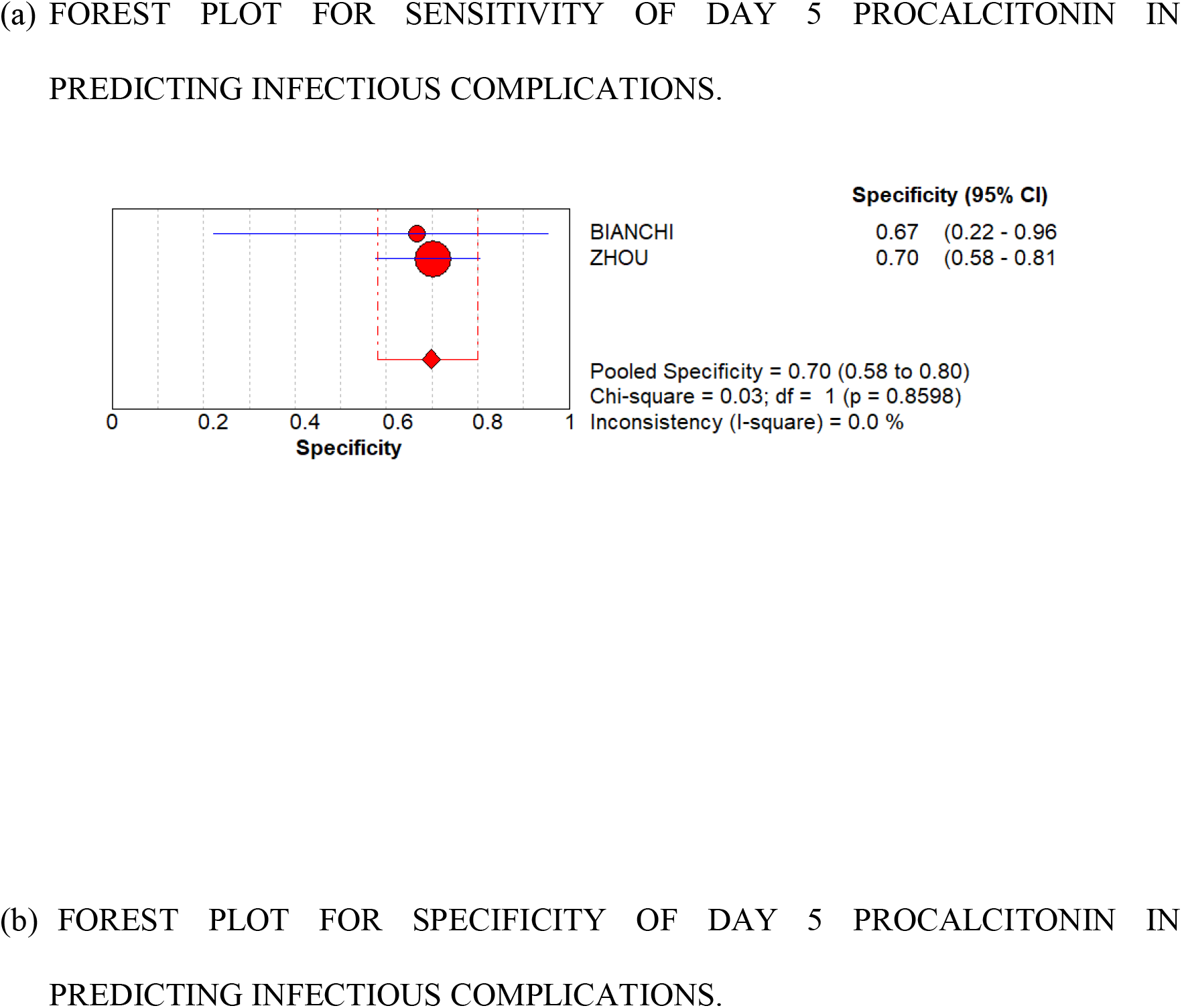
(a) FOREST PLOT FOR SENSITIVITY FOR DAY 5 PCT (b) FOREST PLOT FOR SPECIFICITY FOR DAY 5 PCT.

**FIGURE 8.**
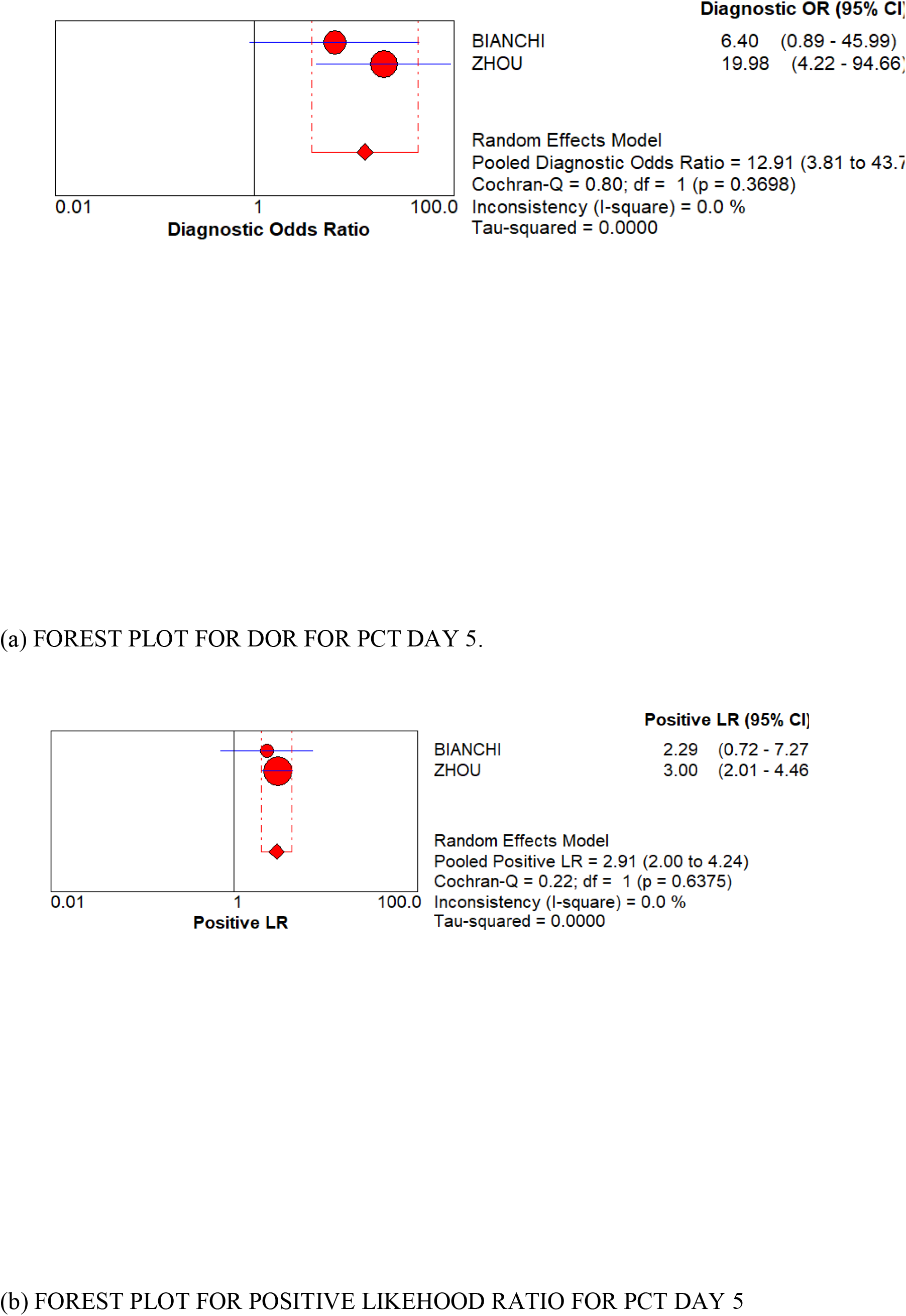
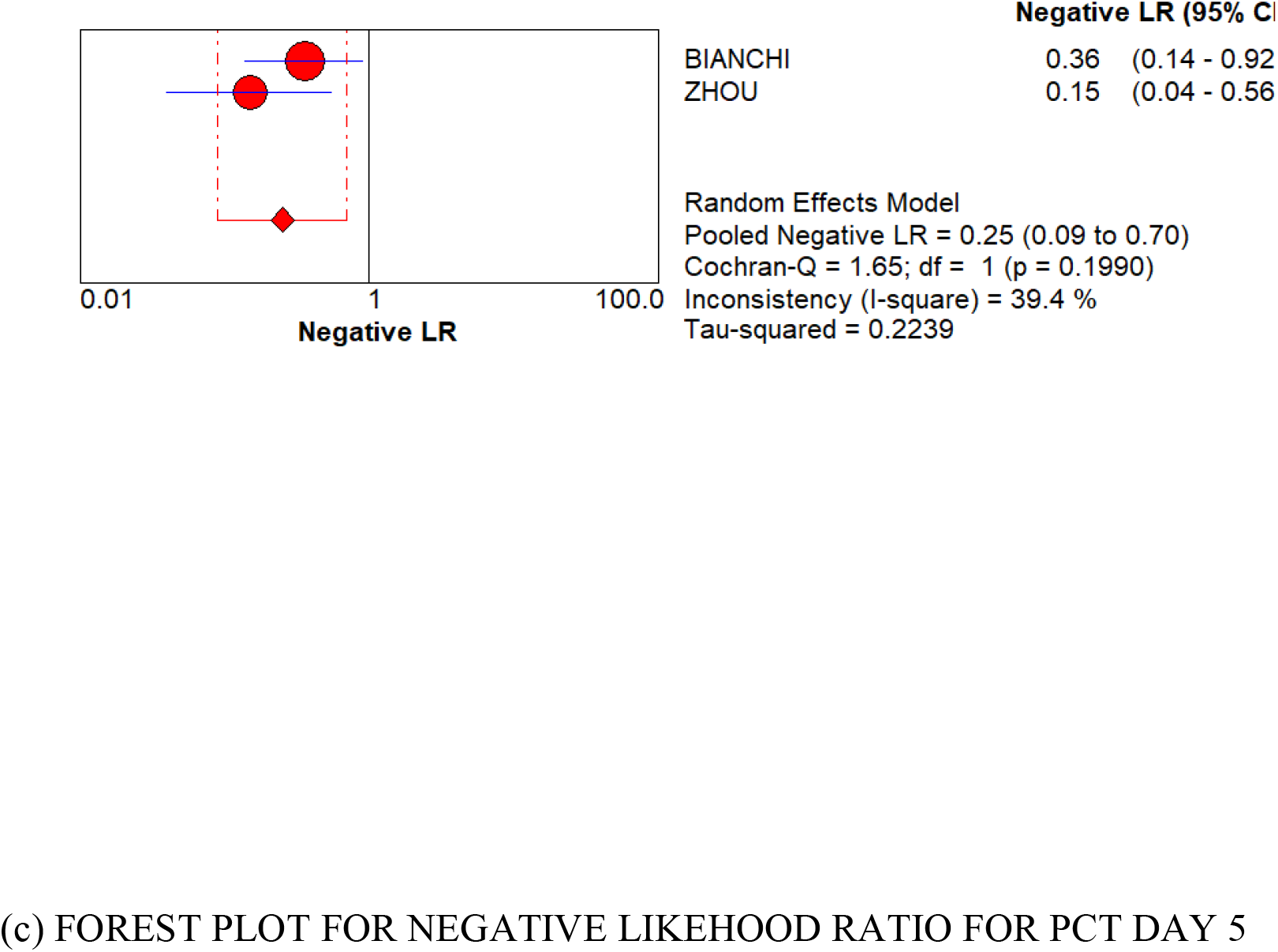
(a) FOREST PLOT FOR DOR FOR PCT DAY 5. (b) FOREST PLOT FOR POSITIVE LIKEHOOD RATIO FOR PCT DAY 5 (c) FOREST PLOT FOR NEGATIVE LIKEHOOD RATIO FOR PCT DAY 5.

**FIGURE 9.**
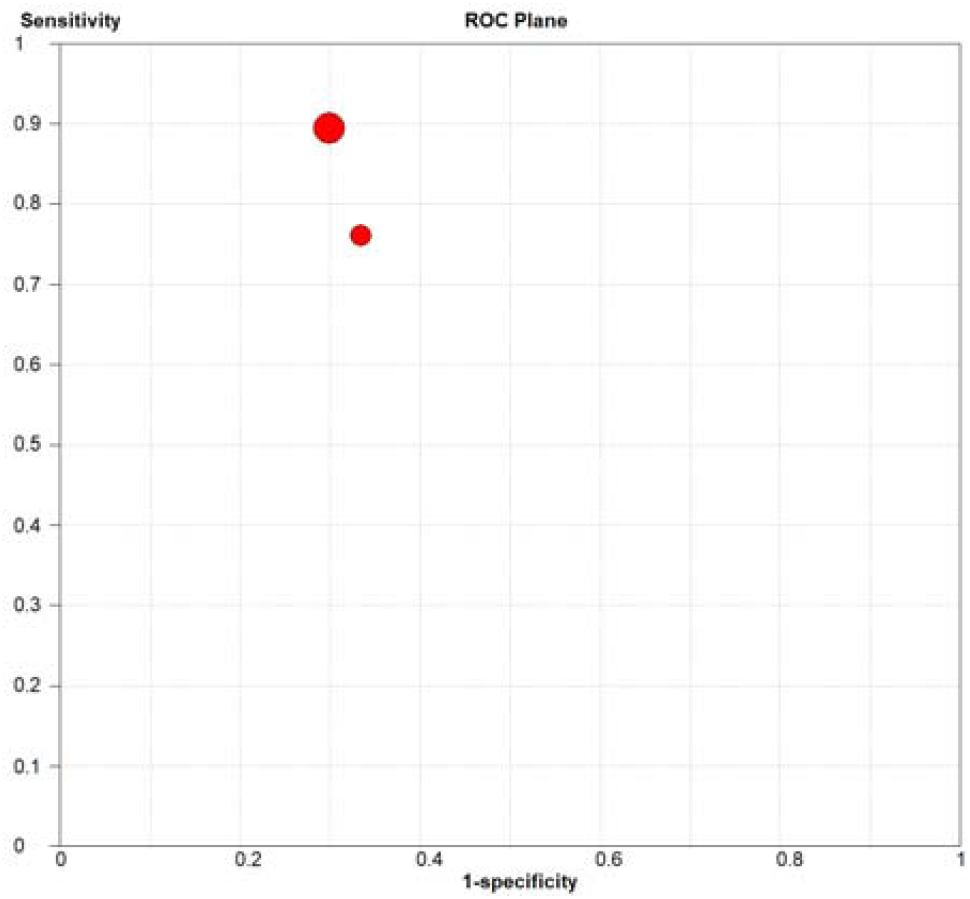
ROC FOR PCT DAY 5.

## DISCUSSION

In our meta-analysis we have shown that post-operative PCT is a good indicator to predict post-operative infectious complications after pancreatic surgeries. Tan et al. [5] and cousin et al. [25] had done similar meta-analysis showing use of PCT as a predictor for infectious complications following colorectal surgeries. However, to our knowledge this is the first meta-analysis showing use fullness of post-operative PCT levels in predicting infectious complications following pancreatic surgeries.

Survival Sepsis Guidelines 2016.[26] suggests use of PCT as a marker for diagnosing sepsis as well as marker for de-escalation of antibiotics and its use in management of sepsis is gaining popularity now. We decided to use PCT levels at day 3 and day 5 as evidences suggests that PCT can be falsely elevated in first 2 post-operative days. [27,28,29].

Many studies also recommend use of C-reactive Protein.(CRP) as a marker of infections in colorectal and pancreatic surgeries [30,31,32]. However, majority of studies showed that highest sensitivity and specificity of CRP in predicting infectious complications is at day 5 which may be too late particularly in Enhanced Recovery After Surgery or ERAS era and also as per survival sepsis guidelines PCT levels can also help in de-escalations of post-operative antibiotics. In our study we have shown that day 3 PCT level can predict post-operative infectious complications with high sensitivity and specificity.

Pooled Area Under Curve of day 3 procalcitonin level was 0.8453, pooled sensitivity 74%,pooled specificity 79%, Diagnostic Odds ratio 11.03, positive like hood ratio 3.17 and negative like hood ratio 0.31 which suggest day 3 Procalcitonin is a very good predictive test for post-operative infectious complications. Youden Index was 0.53 which also confirms above findings.[33].

Sensitivity, specificity, Diagnostic Odds ratio, positive and negative like hood ratio of day 5 procalcitonin levels were 83%, 70%, 12.91,2.91 and 0.25 with Youden index 0.53. However, only two studies evaluated day 5 PCT levels so more evidence are needed to confirm above findings.

There are certain limitations of our analysis. As, limited number of studies available we need more studies and meta-analysis to confirm our findings. Heterogeneity was moderate in some analysis which can be due to differencs in methodology and study design. Another limitation is majority of studies included pancreaticoduodenectomies only so to confirm these findings in distal pancreatectomies including laparoscopic distal pancreatectomies we need more data.

However this is one of the early meta-analysis which confirms day 3 PCT level can predict infectious complications in pancreatic surgeries.

In, Conclusion procalcitonin is an important marker particularly postoperative day 3 pct level can predict infectious complications after pancreatic duodenectomy.

## Data Availability

data will be made available on demand

---

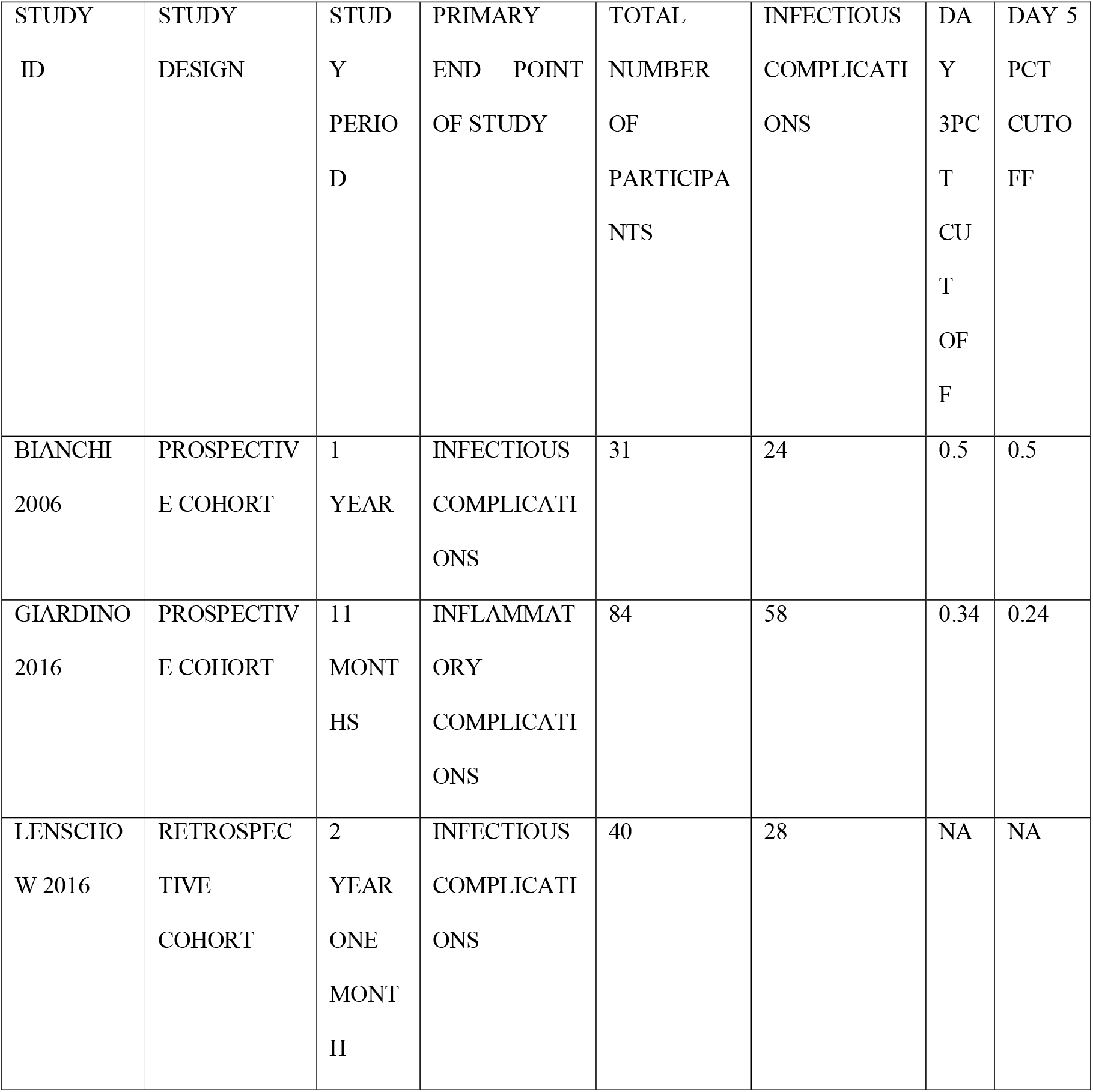
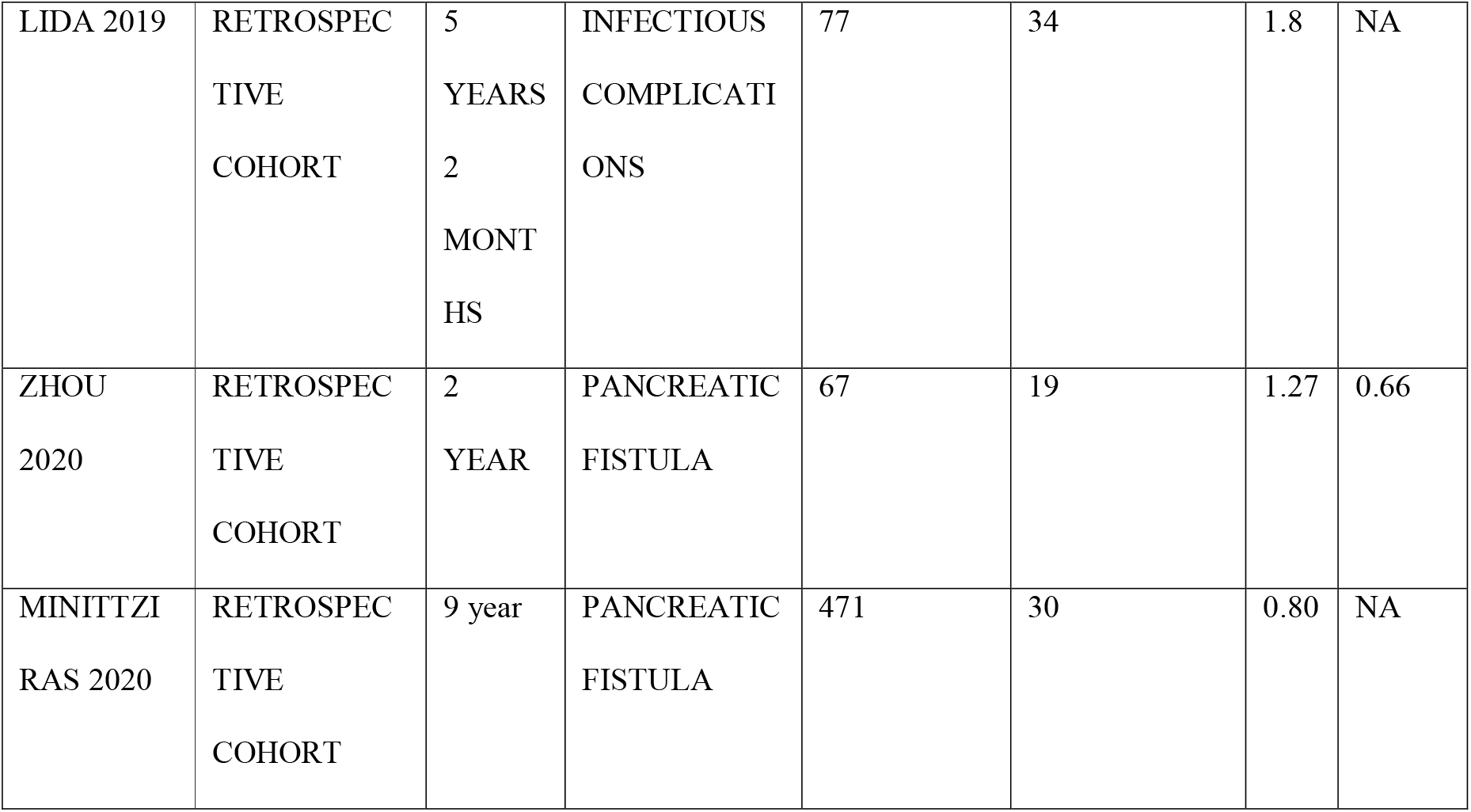

